# Tear-Based Neurodegenerative Biomarkers for Alzheimer’s Disease Screening in Latin America

**DOI:** 10.1101/2025.10.17.25338259

**Authors:** Maisie Bailey, Anshule Takyar, Marleny Nolasco, Richard Trantow, Danilo A Sanchez Coronel, Robert H Gilman, Monica M Diaz

## Abstract

**Background:** Alzheimer’s disease (AD) prevalence is rising globally, with low- and middle-income countries (LMICs) facing diagnostic challenges. We investigated the potential for tear-based biomarkers as a non-invasive diagnostic tool for AD.

**Methods:** Tear samples were collected from 11 patients with MCI or dementia (neurocognitive disorder, NCD) and 5 age-matched cognitively healthy controls at a public hospital in Lima, Peru. Proteins were analyzed for neurodegenerative biomarkers (amyloid-beta 40/42, total tau, phosphorylated tau181 [pTau181], α-synuclein, and neurofilament light).

**Results:** A custom digital ELISA assay on the NanoMosaic platform revealed significantly elevated levels of total tau (p=0.0023) and pTau181 (p=0.0016) in NCD samples compared to controls. Additionally, the pTau181 to total tau ratio was markedly higher in the NCD group (p = 0.007).

**Conclusion:** These findings suggest that tear-based biomarkers, particularly total tau and pTau181, hold promise as a feasible, non-invasive approach for AD screening and diagnosis in resource-limited settings.

## Background

Current Alzheimer’s disease (AD) diagnosis relies on comprehensive clinical assessments and advanced diagnostic methods, such as neuroimaging and cerebrospinal fluid (CSF) biomarker testing^1^. These approaches are often invasive, costly, and largely inaccessible in low- and middle-income countries (LMICs), where healthcare infrastructure is limited^2,3^. In clinical practice, lumbar punctures are commonly used to analyze CSF biomarkers, including amyloid-beta 42/40, total tau, and phosphorylated tau (pTau181)^4^. However, this procedure is invasive, uncomfortable for patients and requires skilled personnel. More recently, plasma p-tau 217 has emerged as a highly sensitive and specific biomarker for AD, yet it remains expensive and largely restricted to high-income countries^5,6^. In many LMICs, including those in Latin America, such assays are either unavailable or limited to research settings^2,3,7^, but plasma p-tau 217 levels have been shown to be significantly associated with AD in a large Peruvian sample^8^. These barriers have spurred growing interest in identifying AD biomarkers from more accessible, non-invasive sources. One study have found that phosphorylated-tau can be detected in salivary samples of patients with AD in Brazil^9^, highlighting the utility of using non-invasive diagnostic biomarkers for AD in Latin America.

Globally, approximately 55 million people are currently living with dementia with nearly 10 million new cases each year, 63% of whom live in low-to-middle income countries (LMICs)^10^. Comorbidities associated with aging, including neurocognitive disorders, are also expected to increase^11^. Dementia poses a significant physical, mental, and economic burden not only for patients, but also for families and communities^10^, highlighting the need to identify dementia early-on in order to modify associated risk factors prior to progressing to dementia. Alzheimer’s disease (AD) is the most common cause of dementia and accounts for 60-70% of all cases^12^. Therefore, early diagnosis of AD is critical to optimize management, start treatment if indicated and improve outcome of patients with AD.

Tear fluid has recently emerged as a potential source of protein-based biomarkers for neurological diseases, offering a promising and less invasive alternative to CSF. The lacrimal gland is connected to the brain through the parasympathetic nervous system, with nerve fibers traveling via the facial nerve to the pterygopalatine ganglion, where they synapse and then reach the lacrimal gland, ultimately stimulating tear production^13^, making tears a possible non-invasive biomarker source from the brain. A 2017 study found that 514 tear proteins, of which 365 were found in other tissues and organs of the body^14^. Some studies have suggested a potential association between tear protein elevations and neurodegenerative disorders, including multiple sclerosis (MS), epilepsy and Friedreich’s ataxia^15,16^. Tears have also emerged as a potential media for the detection of biomarkers for AD recently. Early studies have indicated that proteins such as Total Tau and phosphorylated Tau-181 (pTau181) can be measured in tear fluid, opening new possibilities for disease diagnosis^17–19^. However, not all research has found tear-based biomarkers to be effective for neurological disease detection. One study examining tear proteins in relation to diabetic neuropathy, for example, did not find them to be a sufficient tool for detecting the condition, underscoring the need for further validation and disease-specific research^20^. These studies demonstrate the utility of tears as a potential non-invasive biomarker for detection of other neurodegenerative diseases, such as AD and related dementias. However, these studies did not apply biomarkers in LMIC settings for which less readily available and less sophisticated diagnostic strategies are particularly needed.

Developing a rapid, patient-centered test by collection of tear proteins using Schirmer strips would aid in rapid diagnostics of AD, particularly in resource-limited settings such as in Peru, where CSF biomarkers of AD are limited in the clinical setting. We aimed to determine whether AD proteins can be detected in tears of patients with neurocognitive disorder (MCI or dementia), and whether AD protein levels differed in the neurocognitive disorder group compared with control groups. We hypothesized that NanoMosaic platform would yield the highest levels of neurodegenerative biomarker proteins given it is able to detect protein at very low volumes. These aims would allow us to assess the utility of detecting neurodegenerative biomarkers in people with neurocognitive disorder compared with age- and sex-matched healthy people from a public neurological institute in Lima, Peru.

## Methods

### Study Population

Participants eligible for the study included individuals over 18 years of age with a confirmed diagnosis of MCI or dementia (at any stage, based on the Clinical Dementia Rating (CDR) Scale conducted by their treating neurologist) for six months or more since the study evaluation. These individuals had to be accompanied by a next-of-kin or family caregiver capable of providing consent on their behalf. Additionally, participants could include adults who wear contact lenses infrequently, provided they had not worn them for at least one month prior to the study. Healthy adults without any neurological diseases were also eligible to participate as control subjects.

Individuals with Parkinson’s disease, glaucoma, autoimmune conditions, or other illnesses that could affect tear protein concentrations were excluded from the study. Participants were also excluded if they were currently using medications such as doxycycline, corticosteroid analogues, prostaglandins, or anticholinergic drugs that are known to affect tear production. Additional exclusion criteria included an allergy to fluorescein, pregnancy, breastfeeding, ongoing chemotherapy treatment. Those with recent eye injuries or infections (within the last two weeks) or pre-existing ophthalmologic issues, such as dry eye diagnosed by an ophthalmologist, were also not eligible to participate.

### Study Procedures

Tear samples were collected at the Instituto Nacional de Ciencas Neurologicas (*National Institute of Neurological Sciences*) in Lima, Peru, between October 2019 and March 2020. An eligibility criteria questionnaire was administered. Participant disease status was assessed via an assessment by a neurologist sub-specialized in dementia and neurodegenerative disorders, after which a CDR score classified participants according to the following scale: 0=normal cognition, 0.5=mild cognitive impairment, 1=mild neurocognitive disorder, 2=moderate neurocognitive disorder and 3=severe neurocognitive disorder. The patient’s treating neurologist assigned the CDR based on their clinical evaluation and assessment. The treating neurologist’s CDR score on the day of the visit or within the prior 6 months that the tear samples were obtained was used for our study. We did not assign Alzheimer’s disease diagnoses and instead based our classification of participants on the CDR score. CDR 0.5-3 was considered a neurocognitive disorder (MCI or neurocognitive disorder) and CDR 0 was considered a cognitively healthy control.

In total, there were 35 study participants, of which 26 were individuals with MCI or dementia (CDR >=0.5) and 9 were age-matched controls without a diagnosis of MCI nor dementia (CDR 0). Samples were collected using TearFlo Schirmer test strips^21^ using a previously published method^22^. A strip was inserted into the lower conjunctival sac of the each of the participant’s eyes simultaneously without the use of anesthetic. After five minutes, the strips were carefully removed using gloves, and the tear volumes, measured in millimeters using markings on each Schirmer strip, were recorded. The strips were then air-dried, placed in small plastic bags, and stored at -80C for future processing and analyses.

### Sample Processing

Samples were extracted in a 100 microliter solution of phosphate-buffered saline (PBS), 0.5% Tween20, and Roche cOmplete Protease Inhibitor. The strip was divided into smaller fragments and placed into a 1.5 mL tube. The mixture was gently agitated for 2 hours at 4C, and then later centrifuged at 12000 rpm for 10 minutes. Mixture was then stored at -80C until analysis, and each sample was limited to 2 freeze-thaw cycles.

### Tear Analysis

Total protein content was estimated using a 1:40 dilution Bradford assay, after which nonviable samples (undetectable protein content) were excluded from the study (n=19). The remaining 11 neurocognitive disorder samples and 5 control (controls) samples were then analyzed using commercially available enzyme-linked immunosorbent assay (ELISA) kits (ThermoFisher Human TIMP-4 ELISA Kit, 1:4 dilution; Human Tau (Total) ELISA Kit, 1:4 dilution; Human Tau (Phospho) [pT181] ELISA Kit, 1:4 dilution; Human Amyloid beta 40 ELISA Kit, 1:4 dilution; Human Amyloid beta 42 ELISA Kit, 1:4 dilution; Human alpha Synuclein ELISA Kit, 1:4 dilution).

Subsequently, a ThermoFisher multiplex Luminex assay was used to test the samples for Human TIMP-4, total Tau, pT181, amyloid β40, amyloid β42, and α-synuclein at a 1:2 dilution. The Quanterix Simoa neurofilament light (NfL) assay was used to analyze NfL levels. NanoMosaic NanoPlates were used to detect concentrations of Human TIMP-4, total tau, pT181, amyloid β40, amyloid β42, and α-synuclein at low levels. Antibody pairs for each biomarker were spotted onto the NanoPlates, and completed panels were analyzed using a NanoMosaic Tessie NanoPlate reader. The Quanterix Simoa NfL assay was used to analyze neurofilament light in the samples. For each assay, samples were run in duplicate and randomized, and a standard curve was created to calculate and standardize concentrations **(Table 1)**.

**Table 1.** Biomarkers and respective assays used for analysis.

| **Biomarkers** | **Proteomic Assays Used** |
| --- | --- |
| **Total Tau** | Commercial ELISA, Luminex, NanoMosaic |
| **pTau181** | Commercial ELISA, Luminex, NanoMosaic |
| **Aβ40** | Commercial ELISA, Luminex |
| **Aβ42** | Commercial ELISA, Luminex |
| **α-Synuclein** | Commercial ELISA, Luminex |
| **NfL** | Quanterix |

### Tear Biomarker Concentration Normalization

The concentration of each biomarker in tears collected using Schirmer strips was reported as total recovery in ng/mL or pg/mL. Normalization followed the method described in a previously published protocol^22^. To account for differences in sample volumes collected from each participant on the Schirmer strips, we multiplied the assay’s raw concentration by the total extraction volume (0.1 mL) to obtain the total mass of the extracted sample. This mass was then divided by the volume of the Schirmer strip and multiplied by 1000 to determine the final protein concentration, expressed in pg/mL.

### Ethical Approval

Ethical approval was obtained from the Instituto Nacional de Ciencias Neurologicas Institutional Review Board in Lima, Peru. All participants or their proxy provided written informed consent.

### Statistical Analyses

Descriptive statistics were performed on all participant characteristics and laboratory results. T-tests were performed to determine statistically significant differences between neurocognitive disorder and control groups for all laboratory results. Analyses were performed using STATA 15.0.

## Results

Of the 35 number of available samples, 19 had no detectable protein content, which was determined using a Bradford assay and subsequent Human TIMP-4 ELISA assay. Samples that had no detectable protein concentrations were excluded from further analysis. Therefore, 16 participants’ samples were included in the biomarker analyses. Demographics and medical characteristics of the participants (n=16) are shown in **Table 2**. After concentration normalization, neurocognitive disorder and control group summary statistics (**Table 3**) were compared using a Welch’s t-test (**Figure 1**).

**Figure 1.**
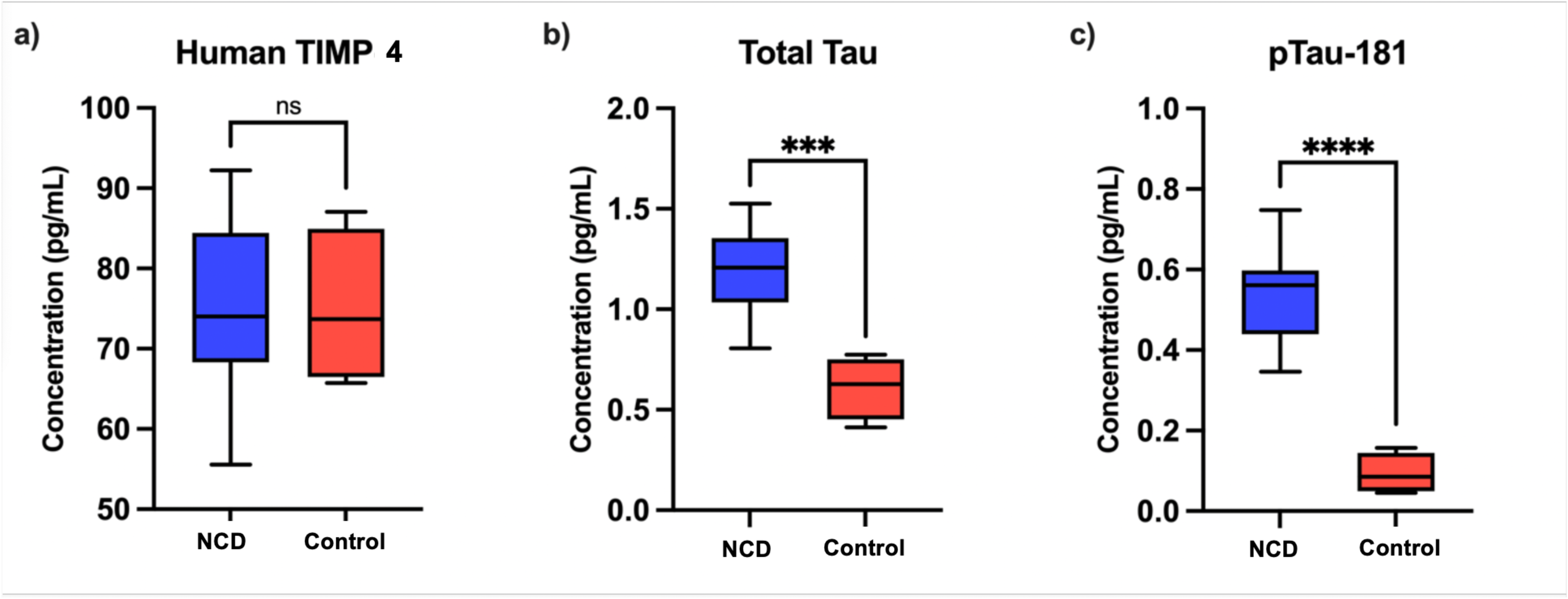
Comparison of biomarker concentrations (pg/mL) by neurocognitive disorder status group. (a) Human TIMP-1 concentrations, included as a negative control, were not significantly different between neurocognitive disorder and control samples (p=0.8887). (b) Total tau concentrations (pg/mL) and (c) pTau181 concentrations (pg/mL) were significantly higher in neurocognitive disorder and control samples (p=0.0006 and p<0.0001 respectively).

**Table 2.**
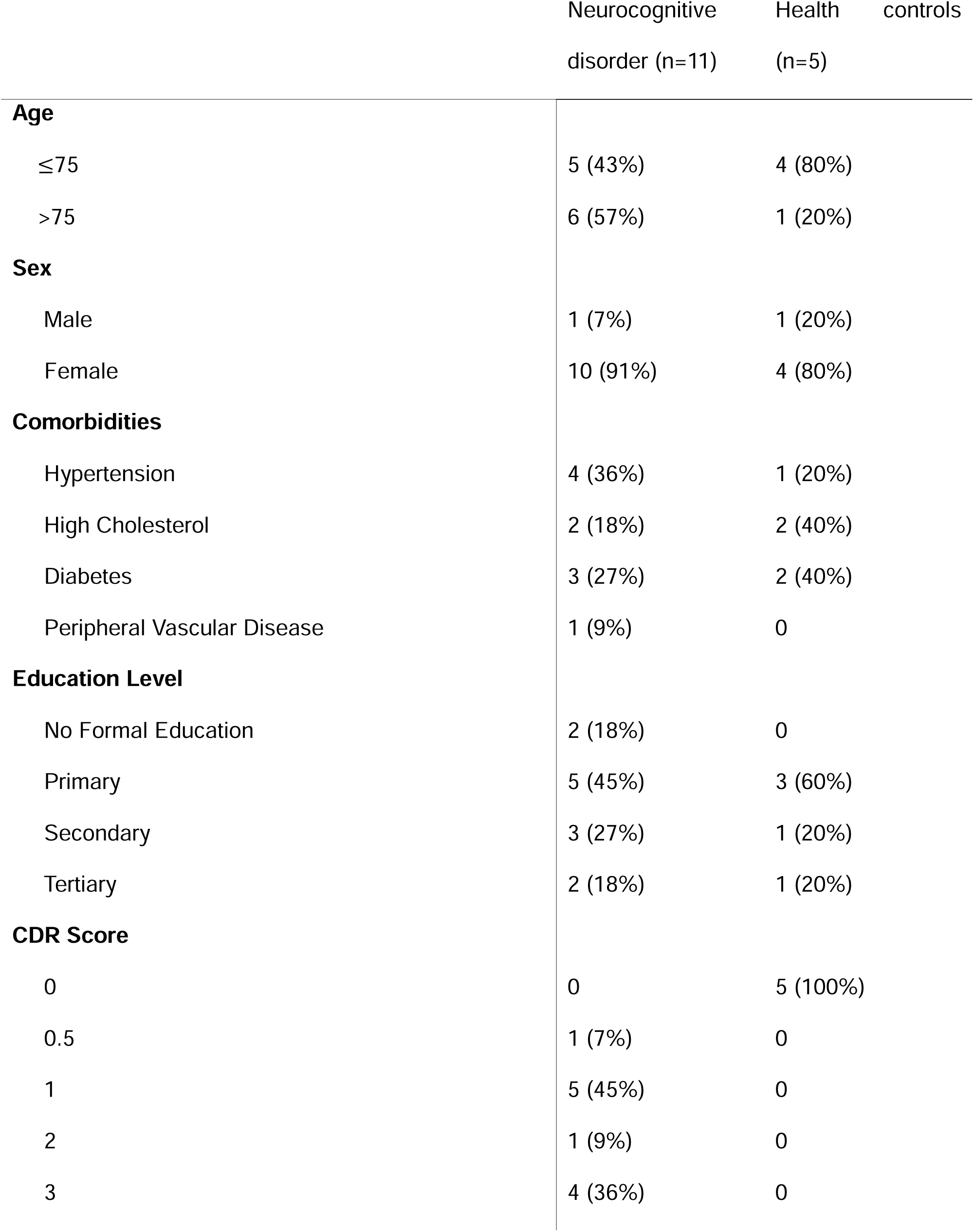
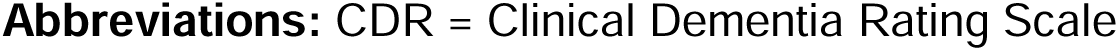
Summary Demographics of Study Participants in Tear Analysis (N=16)

|  | Neurocognitive disorder (n=11) | Health controls (n=5) |
| --- | --- | --- |
| **Age** |  |  |
| ≤75 | 5 (43%) | 4 (80%) |
| >75 | 6 (57%) | 1 (20%) |
| **Sex** |  |  |
| Male | 1 (7%) | 1 (20%) |
| Female | 10 (91%) | 4 (80%) |
| **Comorbidities** |  |  |
| Hypertension | 4 (36%) | 1 (20%) |
| High Cholesterol | 2 (18%) | 2 (40%) |
| Diabetes | 3 (27%) | 2 (40%) |
| Peripheral Vascular Disease | 1 (9%) | 0 |
| **Education Level** |  |  |
| No Formal Education | 2 (18%) | 0 |
| Primary | 5 (45%) | 3 (60%) |
| Secondary | 3 (27%) | 1 (20%) |
| Tertiary | 2 (18%) | 1 (20%) |
| **CDR Score** |  |  |
| 0 | 0 | 5 (100%) |
| 0.5 | 1 (7%) | 0 |
| 1 | 5 (45%) | 0 |
| 2 | 1 (9%) | 0 |
| 3 | 4 (36%) | 0 |

**Abbreviations:** CDR = Clinical Dementia Rating Scale

**Table 3.** Mean biomarker concentrations (pg/mL) and interquartile range for neurocognitive disorder and control groups.

|  | Neurocognitive disorder | Healthy controls |
| --- | --- | --- |
| Human TIMP-4 | 75.20 (67.06-83.34) | 75.05 (59.58-90.52) |
| Total Tau | 1.186 (1.028-1.343) | 0.6104 (0.3627-0.8581) |
| pTau181 | 0.5353 (0.4512-0.6194) | 0.0935 (0.0138-0.1731) |

Abbreviations: = pTau181 = phosphorylated tau 181; TIMP-4 = Tissue inhibitor of metalloproteinases-4

Human TIMP-1 concentrations, included as a laboratory control, were not significantly different between neurocognitive disorder and control samples (p=0.8887). Total tau concentrations (pg/mL) and pTau181 concentrations (pg/mL) were significantly higher in neurocognitive disorder and control samples (p=0.0006 and p<0.0001 respectively); **Figure 2**. pTau181 to total tau ratio for each participant, grouped by neurocognitive disorder status. Neurocognitive disorder samples had significantly higher ratios (p=0.007) than control samples.

**Figure 2.**
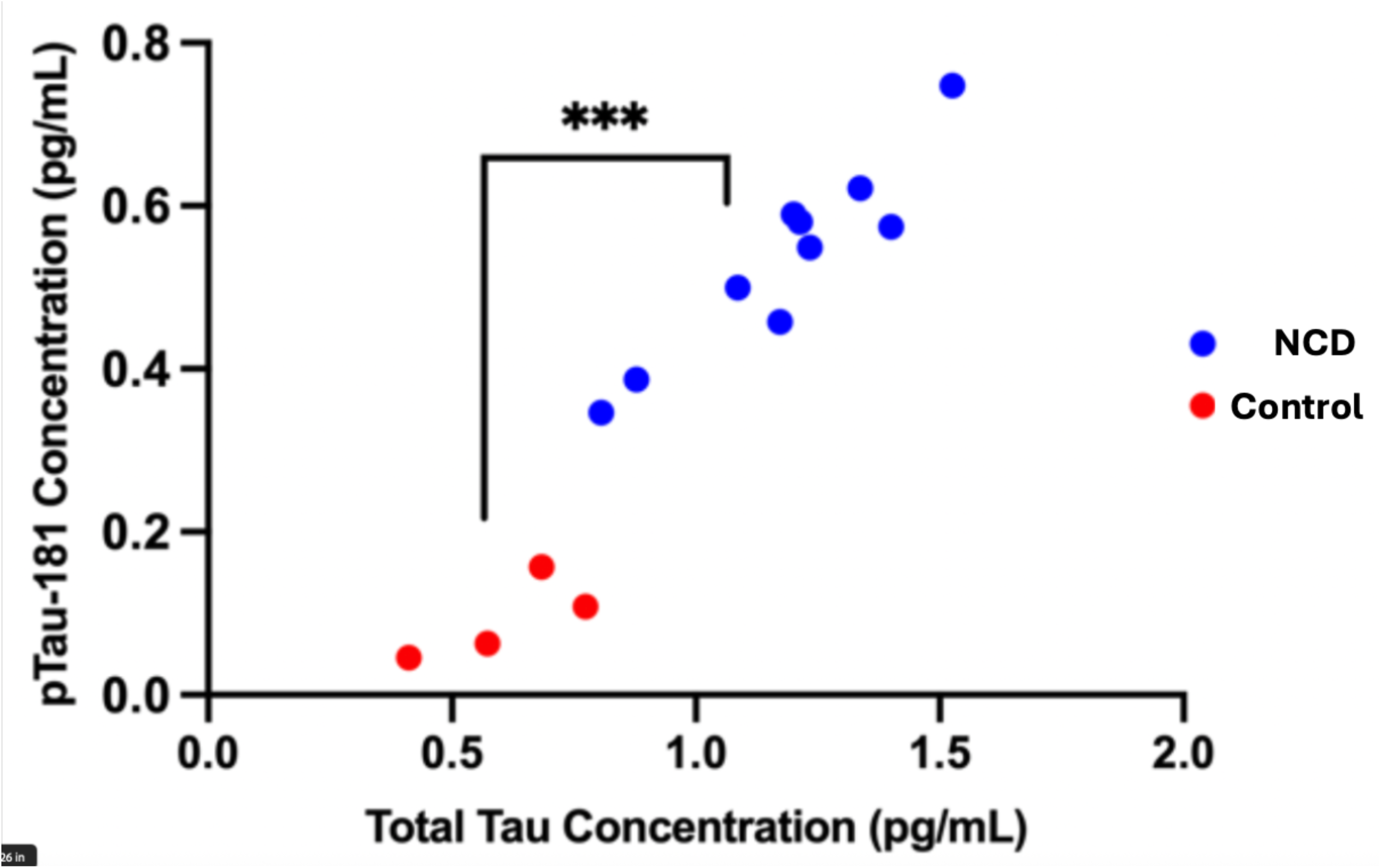
pTau181 to total tau ratio for each participant, grouped by neurocognitive disorder status. pTau181 to total tau ratio for each participant, grouped by disease status. Neurocognitive disorder samples had significantly higher ratios (p=0.007) than control samples.

## Discussion

The increasing prevalence of AD, particularly in LMICs including in Peru, highlights the critical need for accessible and cost-effective diagnostic tools. Our findings demonstrated a significant elevation of total tau and pTau181 levels in the tear fluid of individuals with a CDR score of 1 or greater, which supports the potential role of tears as biomarkers for disease screening in this small sample. Our findings align with previous research showing altered tear protein expression in AD, but extend this prior work by applying high-sensitivity platforms (NanoMosaic, Quanterix) capable of detecting low-abundance proteins in small-volume tear samples. These advances are critical for adapting biomarker detection to non-invasive collection methods such as Schirmer strips.

Previous efforts assessing biomarkers in tear fluid have been hindered by low protein concentrations in collected samples, which makes standard proteomic analysis difficult and imprecise. Point-of-care diagnostics are increasingly becoming necessary to improve access to care in remote or rural areas in LMICs^23^. About 20 million people in Peru live outside of the capital city, most of whom live in rural regions with limited access to neurologists or geriatricians^24^, plasma or CSF collection, lack of laboratory infrastructure, freezer access for storage of plasma samples for diagnosis^25,26^. Moreover, there is widespread stigma associated with lumbar punctures which are not routinely done in clinical care for the diagnosis of AD^7^ in Peru. Developing non-invasive techniques that require little laboratory processing prior to storage is needed to improve access to AD and MCI diagnostics.

Tear proteins may serve as a useful non-invasive adjunct to the current gold standard for dementia diagnosis in LMICs^27^and may help distinguish MCI and AD from healthy controls. Plasma and CSF biomarkers, however, require laboratory processing (centrifuge, aliquoting) limiting utility in LMICs, especially in rural areas. Based on the 2024 National Institute on Aging and the Alzheimer’s Association (NIA-AA) diagnostic criteria for AD, AD is diagnosed by cortical atrophy on MRI, presence of amyloid on amyloid PET scan, and/or altered neurodegenerative biomarker levels in CSF or plasma, including elevated plasma p-tau_217_ ^28^ but these studies are expensive and inaccessible in most LAC countries, including in Peru^29,30^.

Proteins found in tears are also found in other organs and can be used as non-invasive markers of conditions such as AD, multiple sclerosis, Parkinson’s disease, epilepsy and Friedreich’s ataxia^18,19,31–33^. Although plasma biomarkers are now part of the revised 2024 NIA-AA diagnostic criteria for diagnosis of AD^28^ and may be used to stage AD, they still require laboratory processing prior to storage, phlebotomist training and an uncomfortable needlestick. Demonstrating the feasibility of tear proteins to distinguish between AD, MCI and healthy controls is needed, particularly to diagnose MCI early and aid in dementia risk factor modification to prevent disease progression.

Importantly, the potential for tear biomarkers to complement or substitute more invasive methods such as CSF analysis or neuroimaging aligns with the global push toward accessible diagnostics. Early and reliable detection of AD in LMICs could significantly impact clinical outcomes by enabling timely interventions and resource allocation. Despite our findings demonstrating that total tau and pTau181 protein levels in tears are elevated in people with clinical neurocognitive disorder compared with controls, challenges remain. The difficulty in extracting sufficient protein concentrations from tear samples and the statistical insignificance of other biomarkers, such as NfL, emphasize the need for further refinement of sample processing techniques. Additionally, protein concentration was undetectable in n=21 samples, reducing our overall sample size and potentially impacting statistical power. Thus, sample storage and transfer can be improved upon in future studies. Addressing these limitations and optimizing tear sample handling will allow for robust future analyses with currently developing proteomic pipelines.

Future directions should focus on expanding the biomarker panel to include additional proteins implicated in AD progression, validating these findings across larger and more diverse cohorts, and standardizing tear sample collection and processing protocols. Collaborative efforts involving technological innovation, cross-institutional studies, and public health initiatives will be essential to translate these findings into clinical practice. In conclusion, this study contributes to the growing evidence of tear fluid as a feasible medium for biomarker-based AD screening. While challenges remain, the results provide a foundation for the development of a non-invasive, accessible diagnostic tool that could transform AD detection, particularly in under-resourced healthcare systems.

## Data Availability

All data produced in the present study are available upon reasonable request to the authors

## Acknowledgements

We would like to acknowledge the participants that participated in this study.

## Consent Statement

All participants or their proxy provided written informed consent.

## References

1. Atri A, Dickerson BC, Clevenger C, et al. Alzheimer’s Association clinical practice guideline for the Diagnostic Evaluation, Testing, Counseling, and Disclosure of Suspected Alzheimer’s Disease and Related Disorders (DETeCD-ADRD): Executive summary of recommendations for primary care. Alzheimer’s & Dementia. n/a(n/a). doi:10.1002/alz.14333

2. Almubaslat F, Sanchez-Boluarte SS, Diaz MM. A review of neurological health disparities in Peru. Front Public Health. 2023;11:1210238. doi:10.3389/fpubh.2023.1210238

3. Aranda MP, Kremer IN, Hinton L, et al. Impact of dementia: Health disparities, population trends, care interventions, and economic costs. J Am Geriatr Soc. 2021;69(7):1774–1783. doi:10.1111/jgs.17345

4. Janelidze S, Mattsson N, Palmqvist S, et al. Plasma P-tau181 in Alzheimer’s disease: relationship to other biomarkers, differential diagnosis, neuropathology and longitudinal progression to Alzheimer’s dementia. Nat Med. 2020;26(3):379–386. doi:10.1038/s41591-020-0755-1

5. Mattsson-Carlgren N, Janelidze S, Palmqvist S, et al. Longitudinal plasma p-tau217 is increased in early stages of Alzheimer’s disease. Brain. 2020;143(11):3234–3241. doi:10.1093/brain/awaa286

6. Janelidze S, Stomrud E, Smith R, et al. Cerebrospinal fluid p-tau217 performs better than p-tau181 as a biomarker of Alzheimer’s disease. Nat Commun. 2020;11:1683. doi:10.1038/s41467-020-15436-0

7. Saylor D, Elafros M, Bearden D, et al. Patient, Provider, and Health Systems Factors Leading to Lumbar Puncture Nonperformance in Zambia: A Qualitative Investigation of the “Tap Gap.” Am J Trop Med Hyg. 2023;108(5):1052–1062. doi:10.4269/ajtmh.22-0699

8. Pandey N, Yang Z, Cieza B, et al. Plasma phospho-tau217 as a predictive biomarker for Alzheimer’s disease in a large south American cohort. Alzheimers Res Ther. 2025;17(1):1. doi:10.1186/s13195-024-01655-w

9. Dos Santos GAA, do Vale F de AC, Fazan VPS. Prospecting salivary tau as a diagnostic for Alzheimer’s type dementia. Dement Neuropsychol. 2025;19:e20240253. doi:10.1590/19805764-DN-2024-0253

10. Prince M, Guerchet M, Prina M. The Epidemiology and Impact of Dementia - Current State and Future Trends. WHO Thematic Briefing. Published online 2022. Accessed April 21, 2025. https://hal.science/hal-03517019

11. Estimation of the global prevalence of dementia in 2019 and forecasted prevalence in 2050: an analysis for the Global Burden of Disease Study 2019. Lancet Public Health. 2022;7(2):e105–e125. doi:10.1016/S2468-2667(21)00249-8

12. Dementia. Accessed April 21, 2025. https://www.who.int/news-room/fact-sheets/detail/dementia

13. Dartt DA. Neural Regulation of Lacrimal Gland Secretory Processes: Relevance in Dry Eye Diseases. Prog Retin Eye Res. 2009;28(3):155–177. doi:10.1016/j.preteyeres.2009.04.003

14. Qin W, Zhao C, Zhang L, Wang T, Gao Y. A Dry Method for Preserving Tear Protein Samples. Biopreservation and Biobanking. 2017;15(5):417–421. doi:10.1089/bio.2016.0117

15. Örnek N, Dağ E, Örnek K. Corneal sensitivity and tear function in neurodegenerative diseases. Curr Eye Res. 2015;40(4):423–428. doi:10.3109/02713683.2014.930154

16. Markoulli M, You J, Kim J, et al. Corneal Nerve Morphology and Tear Film Substance P in Diabetes. Optometry and Vision Science. 2017;94(7):726. doi:10.1097/OPX.0000000000001096

17. Van De Sande N, Ramakers IHGB, Visser PJ, et al. Tear biomarkers for Alzheimer’s disease screening and diagnosis (the TearAD study): design and rationale of an observational longitudinal multicenter study. BMC Neurol. 2023;23(1):293. doi:10.1186/s12883-023-03335-y

18. Kalló G, Emri M, Varga Z, et al. Changes in the Chemical Barrier Composition of Tears in Alzheimer’s Disease Reveal Potential Tear Diagnostic Biomarkers. PLOS ONE. 2016;11(6):e0158000. doi:10.1371/journal.pone.0158000

19. Naseri N, Ahmadi N, Harirchian MH, Asadi A, Ghafari V, Rostami M. The Importance of Proteomics in Saliva and Tears as Potential Non-invasive Methods for Identifying Biomarkers in the Prognosis and Diagnosis of Multiple Sclerosis Patients. Arch Neurosci. 2024;11(4). doi:10.5812/ans-145578

20. Iyengar MF, Soto LF, Requena D, et al. Tear biomarkers and corneal sensitivity as an indicator of neuropathy in type 2 diabetes. Diabetes Res Clin Pract. 2020;163:108143. doi:10.1016/j.diabres.2020.108143

21. HUB Pharmaceuticals. TearFlo Schirmer test strips. https://hubrx.com/our-products/tearflo/.

22. VanDerMeid KR, Su SP, Krenzer KL, Ward KW, Zhang JZ. A method to extract cytokines and matrix metalloproteinases from Schirmer strips and analyze using Luminex. Mol Vis. 2011;17:1056–1063. Accessed April 21, 2025. https://www.ncbi.nlm.nih.gov/pmc/articles/PMC3086627/

23. Thwala LN, Ndlovu SC, Mpofu KT, Lugongolo MY, Mthunzi-Kufa P. Nanotechnology-Based Diagnostics for Diseases Prevalent in Developing Countries: Current Advances in Point-of-Care Tests. Nanomaterials (Basel). 2023;13(7):1247. doi:10.3390/nano13071247

24. “Hay solo 157 geriatras para 3 millones de adultos mayores”. El Comercio. https://elcomercio.pe/lima/hay-157-geriatras-3-millones-adultos-mayores-335785-noticia/.

25. Parra MA, Baez S, Sedeño L, et al. Dementia in Latin America: Paving the way toward a regional action plan. Alzheimers Dement. 2021;17(2):295–313. doi:10.1002/alz.12202

26. Ibanez A, Parra MA, Butler C, Latin America and the Caribbean Consortium on Dementia (LAC-CD). The Latin America and the Caribbean Consortium on Dementia (LAC-CD): From Networking to Research to Implementation Science. J Alzheimers Dis. 2021;82(s1):S379–S394. doi:10.3233/JAD-201384

27. Sachdev PS, Blacker D, Blazer DG, et al. Classifying neurocognitive disorders: the DSM-5 approach. Nat Rev Neurol. 2014;10(11):634–642. doi:10.1038/nrneurol.2014.181

28. Jack CRJ, Andrews JS, Beach TG, et al. Revised criteria for diagnosis and staging of Alzheimer’s disease: Alzheimer’s Association Workgroup. Alzheimers Dement. 2024;20(8):5143–5169. doi:10.1002/alz.13859

29. Parra MA, Baez S, Allegri R, et al. Dementia in Latin America: Assessing the present and envisioning the future. Neurology. 2018;90(5):222–231. doi:10.1212/WNL.0000000000004897

30. Lopera F, Custodio N, Rico-Restrepo M, et al. A task force for diagnosis and treatment of people with Alzheimer’s disease in Latin America. Front Neurol. 2023;14:1198869. doi:10.3389/fneur.2023.1198869

31. Lew M, Janga S, Ju Y, et al. Biomarkers for Parkinson’s Disease with Reflex Tears Stratified by Disease Duration (S16.002). Neurology. 2022;98(18_supplement):3750. doi:10.1212/WNL.98.18_supplement.3750

32. Król-Grzymała A, Sienkiewicz-Szłapka E, Fiedorowicz E, Rozmus D, Cieślińska A, Grzybowski A. Tear Biomarkers in Alzheimer’s and Parkinson’s Diseases, and Multiple Sclerosis: Implications for Diagnosis (Systematic Review). International Journal of Molecular Sciences. 2022;23(17):10123. doi:10.3390/ijms231710123

33. Kaštelan S, Braš M, Pjevač N, et al. Tear Biomarkers and Alzheimer’s Disease. Int J Mol Sci. 2023;24(17):13429. doi:10.3390/ijms241713429

